# Genetic relatedness of Cambodian *Plasmodium falciparum* isolates is driven by geography and occupation

**DOI:** 10.64898/2026.01.29.26344731

**Authors:** Emma Rowley, Bing Guo, Mariusz Wojnarski, Michele D. Spring, Sabaithip Sriwichai, Brian A. Vesely, Joana C. Silva, Norman C. Waters, Sasikanya Thaloengsok, Piyaporn Saingam, Chaiyaporn Chaisatit, Paphavee Lertsethtakarn, Somethy Sok, Satharath Prom, Sidhartha Chaudhury, Chanthap Lon, Dysoley Lek, David Saunders, Timothy D. O’Connor, Shannon Takala-Harrison

## Abstract

**Background:** Cambodia is nearing *Plasmodium falciparum* elimination, but targeted interventions are needed to prevent resurgence. Identity-by-descent (IBD)-based methods offer insight into parasite relatedness and transmission dynamics.

**Methods:** We estimated pairwise genetic relatedness between 107 *P. falciparum* isolates collected from military personnel and farmers in Cambodia between 2014 and 2016. All isolates underwent whole genome sequencing. Relatedness was defined as the proportion of the genome shared IBD between isolates. We evaluated associations between relatedness and epidemiologic factors, including occupation, location, time, age, and sex. Infomap community detection was used to identify clusters of highly related parasites and their association with covariates.

**Results:** Mean IBD sharing was high (40%), with greater relatedness among isolates from Oddar Meanchey than from Kratie province (p<2.2e-16). Within Oddar Meanchey, isolates from individuals with different occupations shared 7.2% less of their genome IBD than farmer pairs (probability of direction (pd)=99.9), after adjusting for age. Highly related clusters (>90% IBD) were associated with occupation, K13 mutation (C580Y vs. R539T), and PfCRT mutation (p= 0.0025, 0.0001, and 0.0001, respectively). Isolates from mixed-occupation pairs had lower odds of high relatedness (>90% IBD) than pairs from farmers (pd=0.958).

**Conclusions:** High overall IBD sharing may reflect a combination of recent elimination efforts and expansion of drug-resistant lineages. Substantial differences in relatedness between provinces underscore the role of geography in shaping relatedness patterns. Lower relatedness among mixed-occupation pairs compared to farmer pairs suggests potential occupation-specific transmission sources and supports tailoring interventions to local micro-epidemiologic factors.

## Background

*Plasmodium* parasites cause malaria, a vector-borne disease transmitted by *Anopheles* mosquitoes. Over the past two decades, the Greater Mekong Subregion (GMS) has made substantial progress towards malaria elimination, with hard-to-reach populations now representing the primary barrier to complete elimination.^1^ Artemisinin resistance, first detected in western Cambodia, emerged and spread throughout the GMS, fueling urgency to eliminate malaria in the region.^1^ Cambodia has reached historically low levels of *Plasmodium falciparum* malaria, with remaining cases during this elimination period mostly affecting high-risk groups: forest goers, outdoor workers, mobile and migrant populations, border residents, and working-age males.^2–4^ Identifying infection sources and risk factors in these populations is essential to prevent resurgence.

Military personnel and farmers represent key high-risk groups. Military personnel are high risk due to high mobility and frequent deployment in historically high-prevalence border regions.^5,6^ Data on malaria infections in military populations is sparse; however previous literature has noted higher odds of infection compared to other occupations.^5,7^ Farmers also face increased risk due to prolonged outdoor exposure, particularly during harvest seasons.^8,9^ While both occupations are high-risk, their exposure patterns may differ.

As malaria transmission declines, targeted elimination strategies are needed to address remaining pockets of transmission.^10^ One approach is to identify common sources of infection and define discrete clusters of cases with shared risk factors or exposure histories for intervention. While epidemiologic surveys and travel histories offer valuable insights, genetic relatedness between parasites provides a direct measure of infection ancestry allowing identification of shared infection sources.^10^ Identity-by-descent (IBD) analysis identifies regions of the genome inherited from a common ancestor, with closely related parasites sharing long IBD segments that break down over generations due to recombination.^11^ In *Plasmodium*, IBD analysis has shown greater resolution than non-recombination-based methods.^12–16^

This study investigated the association between occupation, as a military member or farmer, and parasite relatedness measured via IBD while considering potential confounders, including time and location. We hypothesized that military personnel and farmers would acquire infections from distinct sources, leading to higher relatedness within each group than between groups. By integrating IBD analysis with epidemiologic metadata, this study provides insights into whether military personnel and farmers acquire infections from distinct transmission sources, information critical for tailoring elimination strategies.

## Methods

### Study Population and Data Collection

This cross-sectional study used samples and survey data from two prior randomized clinical trials in Cambodia. The first trial (2014–2015) in Oddar Meanchey and Kratie provinces compared atovaquone-proguanil vs. artesunate-atovaquone-proguanil for the treatment of uncomplicated *P. falciparum* malaria. Eligible participants were ages 18–65 with parasitemia 100–200,000 parasites/μL. Exclusion criteria included severe malaria, drug contraindications, recent antimalarial use, pregnancy, or lactation.^17^ The original trial was registered with ClinicalTrials.gov (NCT0229747).

The second was a 2016 cluster-randomized controlled open-label pilot study comparing malaria elimination strategies in the Cambodian military in Oddar Meanchey province. Clusters defined by military encampments were randomized. Clusters received either monthly malaria prophylaxis with 3 days of treatment with dihydroartemisinin-piperaquine plus a weekly dose of primaquine or focused screening and treatment with artesunate-mefloquine if confirmed malaria positive. Participants were additionally sub-randomized to receive either insecticide or sham-treated clothing. The study enrolled military volunteers (ages 18–65), their civilian dependents (ages ≥2), and eligible Cambodians in close proximity. Inclusion criteria included informed consent, residing in the study area, prophylaxis adherence, availability for six-month study follow-up, agreement not to seek outside medical care for febrile illnesses, and authorization by local commander to participate in the study if on active duty. Exclusion criteria included medication contraindications, pregnancy, and unwillingness to use contraception.^18,19^ For both studies, all participants provided age, sex, and occupation, and sample collection date and province were recorded. The study was registered on Clinicaltrials.gov (NCT02653898).

### Genomic Data Analysis

*P. falciparum* whole genome sequence data were generated from blood samples collected at diagnosis, before treatment. Samples were leukocyte-depleted to reduce human DNA. DNA was extracted using a Qiagen midi kit and sequenced on an Illumina HiSeq 4000 (150 bp paired end reads). Only *P. falciparum* mono-infections (i.e., no other *Plasmodium* species detected) were retained. Sequencing was approved by the University of Maryland Baltimore IRB, and the sequence data are publicly available (NCBI SRA access number PRJNA1004408).^14^

Single-nucleotide polymorphisms (SNPs) were called using the Genomic Analysis Toolkit’s (GATK) (v4.2.2) best practices and filtered using VQSR.^14,20,21^ Individual sites per genome with <5 reads were masked and indels and non-biallelic SNPs were removed.^14^ Only core-genome SNPs with <20% missing data and minor allele frequency (MAF) >1% were retained, and samples missing >30% of sites were removed using BCFtools (v1.17).^22^ The filtered dataset included 132 isolates.

*F*_*ws*_, a measure of within-host inbreeding that quantifies the proportion of the dominant genotype relative to overall diversity in an infection, was calculated using moimix (v0.0.2).^23^ Isolates with an *F*_*ws*_ >0.95 were considered monoclonal. DEploid (v0.6-beta) was run on 97 monoclonal study isolates and 128 publicly available Cambodian isolates (Pf6) to identify predominant clones.^21,24^ The predominant clones were used as the haplotype panel for DEploidIBD, which was used to deconvolute polyclonal isolates and infer haplotype proportions.^25^ Two isolates were identified as monoclonal by *F*_*ws*_, but polyclonal by DEploid so were included with the polyclonal isolates for deconvolution. DEploidIBD was run five times per isolate at different random seeds, and the run with the highest IBD best-path log-likelihood was used. The predominant haplotype was included in the final dataset if the dominant clone in that run had a proportion ≥0.70 and was at least three times larger than the second-largest clone. The final set included 107 isolates with 10,742 variants: 95 monoclonal and 12 polyclonal infections. Missing sites were imputed with Beagle (v.5.4), assuming a constant recombination rate of 9.6 kb per cM.^26,27^

### Outcome

The outcome of interest was parasite genetic relatedness, defined as the proportion of the genome shared IBD between isolate pairs. IBD was inferred using *hmmIBD* (v2.0.4), software tailored for *Plasmodium*.^28^ The fraction of sites IBD output by *hmmIBD* was used as the proportion of the genome shared IBD. IBD was calculated for all pairwise isolate comparisons (N=5,671), and these comparisons were the unit of analysis.

### Primary Predictor and Covariates

The primary predictor of interest was participant occupation. Although the original study populations included multiple occupations, all isolates with genomic data meeting quality filters came from either military personnel or farmers. Consequently, occupation was analyzed as a binary variable. Additional covariates included sample collection date, province, participant age, sex, and symptom status. Collection dates were grouped into three transmission seasons reflecting natural peaks in the number of samples collected during the study period. These seasons were Season 1 (Dec 2014 – Apr 15, 2015), Season 2 (Apr 16, 2015 – Sep 2015), and Season 3 (Jan – Jun 2016). The sampling gap at the end of 2015 reflects the period between study sampling, not an underlying change in epidemiology.

Since the unit of analysis was each pair of isolates, predictor variables were grouped according to pair characteristics, as follows: occupation (both military personnel, both farmers, or different occupations), collection province (both from Oddar Meanchey, both from Kratie, or from different provinces), sex (both male, both female, or different sexes), time between sample collection (in days), and the age difference between participants (in years).

### Statistical Analyses

Beta regression (brms v2.22.0 in R v4.3.3) was used to model the proportion of the genome shared IBD.^29–31^ Exact 0 and 1 values were adjusted using a small offset (ε=10^-6^). A multiple membership random effect was specified for both samples in each pair to account for the same isolate appearing in multiple pairs. Six MCMC chains were run for 6000 iterations (2000 warm-up), and convergence was confirmed (R-hat<1.01). Variables were included in the final model if associated with mean IBD in bivariate analyses (p<0.05) and had a probability of direction (pd)>0.95, estimated using bayestestR (v0.15.2).^32^ Pd reflects the posterior certainty in the direction of an effect, and effects in the final model with Pd>0.95 were interpreted as having strong evidence of the association. The 95% credible interval (CrI), estimated with bayestestR (v0.15.2), was used to describe the precision and magnitude of the effect.

The Infomap community detection algorithm (igraph v0.11.9) was used to identify clusters of related parasites, applying relatedness thresholds ranging from 10 to 99% IBD, retaining only edges above each cutoff.^33,34^ Varying thresholds served as a sensitivity analysis to test the robustness of associations between cluster membership and epidemiologic predictors. The association between cluster membership and variables of interest was tested using Fisher’s Exact Test with Monte Carlo approximation. Logistic regression models with a multiple membership random effect were also fit with brms to estimate associations.

### Determination of Drug Resistance Markers

Cluster membership’s association with validated markers of piperaquine and artemisinin resistance was assessed. During the sampling timeframe, dihydroartemisinin-piperaquine was the first-line treatment for *P. falciparum* in Cambodia.^3^ Markers considered included mutations in K13 (encoded by the *kelch13* gene, PF3D7_1343700), the *P. falciparum* Chloroquine Resistance Transporter (PfCRT) (encoded by *pfcrt*, PF3D7_0709000), and plasmepsin 2 (*pfpm2*) gene copy number.^35–38^ *Pfpm2* copy number was evaluated using a SYBR Green-based quantitative polymerase chain reaction assay.^39^ The association between cluster membership and drug resistance markers was estimated using Fisher’s Exact Test with Monte Carlo approximation.

## Results

### Polyclonality

Of 313 samples from the original studies, whole genome sequence data was available for 160 *P. falciparum* isolates. Quality filtering removed 28 samples with insufficient genome coverage. Of 132 isolates retained after quality filtering, 97 were defined as monoclonal by *F*_*ws*_ ≥ 0.95, yielding a polyclonality rate of 26.5%. Two of these 97 were called polyclonal by DEploid and treated as polyclonal for IBD analyses. Isolates were retained for IBD analyses if called monoclonal by DEploid or polyclonal with a predominant clone using DEploidIBD, yielding 107 isolates (95 monoclonal + 12 polyclonal).

### Distribution of Variables

Among the 107 isolates **(Supplementary Table 1)**, the average proportion of the genome shared IBD was 0.402 (95% CI: 0.394, 0.410) and ranged from 0 to 1 **(Supplementary Figure 1)**. Descriptive statistics for these isolates are provided in (**Supplementary Table 2)** and were generally representative of the larger set of 313 samples from both studies **(Supplementary Table 3)**. IBD was calculated for all pairwise comparisons among the 107 isolates (N=5,671 pairs). Pairs of isolates collected in Oddar Meanchey shared >20% more of their genome IBD on average compared to Kratie pairs (p<2.2e-16) **(Table 1)**. Isolates from farmers shared ∼6% less of their genome IBD on average than isolates from military members (p=2.68e-05). Pairs of isolates from females were more highly related than pairs from males. Only ten isolate pairs were from female participants, consistent with local epidemiology, where malaria infections occur predominantly in working-age men.^3^ On average, isolates were collected 164.5 days apart, and participants differed in age by 11.4 years. The difference in collection days between isolates and age between individuals were associated with IBD sharing (p=0.0036 and p=0.0169, respectively).

**Table 1.** Descriptive characteristics and IBD sharing across all isolate pairs.

|  | n (%) | Mean IBD | 95% CI | p-value |
| --- | --- | --- | --- | --- |
| **Occupation** |  |  |  | 2.68e-05* |
| Farmer, Farmer | 2415 (42.6) | 0.390 | (0.377, 0.402) |  |
| Farmer, Military | 2590 (45.6) | 0.400 | (0.389, 0.411) |  |
| Military, Military | 666 (11.7) | 0.451 | (0.428, 0.473) |  |
| **Province** |  |  |  | < 2.2e-16* |
| KT, KT | 325 (5.8) | 0.281 | (0.249, 0.312) |  |
| OM, KT | 2106 (37.1) | 0.289 | (0.279, 0.300) |  |
| OM, OM | 3240 (57.1) | 0.487 | (0.477, 0.497) |  |
| **Sex** |  |  |  |  |
| Male, Male | 5151 (90.8) | 0.394 | (0.386, 0.402) | 5.57e-12* |
| Female, Male | 510 (9.0) | 0.476 | (0.452, 0.500) |  |
| Female, Female | 10 (0.2) | 0.544 | (0.431, 0.658) |  |
| **Days between collection** | 5671 (100) | 164.5** | (161.2, 167.7) | 0.0036^ |
| **Age difference (years)** | 5671 (100) | 11.4** | (11.2, 11.6) | 0.0169^ |

\* p-values calculated based on Kruskal-Wallis test. \*\* For continuous variables, mean per pair is reported. ^ p-values calculated based on Spearman's correlation coefficient.

No isolates were collected from military members in Kratie or from female military members leading to sparsity across these levels when stratifying by occupation **(Supplementary Table 5)**. Because occupation was a primary predictor and both occupations were only represented in Oddar Meanchey, downstream regression analyses were restricted to Oddar Meanchey isolates to limit province-related confounding. Similarly, sex was excluded from regression models. Isolate pairs from farmers were sampled 77 days further apart, on average, compared to military pairs (p<2.2e-16).

### Association between predictors and relatedness

Within Oddar Meanchey, occupation had the strongest bivariate association with IBD sharing (p=5.74e-12, **Supplementary Table 5**). Age difference was associated with relatedness (p=0.008), whereas the number of days between collection was not (p=0.485) (**Supplementary Table 5**). Average age differed significantly between farmers and military participants (mean 31.2 vs. 36.9 years, respectively; Welch t-test p=0.0086), so age was considered as a possible confounder.

In multivariable models, occupation remained the strongest predictor of relatedness. Parasites sampled between farmers shared on average 7.2% more of their genome IBD than parasites from different occupational groups, with strong posterior evidence (pd=99.9) (**Table 2**). Each year of age difference was associated with a small decrease in IBD sharing (0.2%) (pd=98.0). The association between collection study and parasite relatedness had low posterior support in multivariable models and therefore was not included in the final model (**Supplementary Table 6E**). Since a subset of isolates from the second study were from asymptomatic cases, symptom status was evaluated as a potential confounder but was not included due to low posterior support (**Supplementary Table 6F&G**).

**Table 2.** Beta regression comparing the proportion of the genome shared IBD between occupation groups adjusted for age difference. R^2^ = 0.172.

|  | Parameter estimates | Marginal change in proportion IBD | 95% CrI* | Pd** |
| --- | --- | --- | --- | --- |
| **Occupation** |  |  |  |  |
| Farmer, Farmer | 0.79 | 0.671 | (0.628, 0.740) |  |
| Military, Farmer | -0.31 | -0.072 | (-0.116,-0.026) | 99.9 |
| Military, Military | -0.23 | -0.053 | (-0.140, 0.031) | 89.3 |
| **Age Difference (1 yr)** | -0.01 | -0.002 | (-0.003,0.000) | 98.0 |

\*Credible interval. \*\*Probability of direction.

### Network analysis

Isolates from Oddar Meanchey shared, on average, 48.7% of their genome IBD. Seven highly related clusters (isolates sharing >90% of their genome IBD) were identified, ranging in size from 2 to 22 isolates (**Figure 1**). Membership in these clusters varied by occupation, K13 mutations, and PfCRT mutations (p=0.0025, p=0.0001, p=0.0001, respectively). *Pfpm2* copy number and transmission season were not associated with cluster membership. One cluster of seven isolates all from military members harbored the K13,R539T mutation and PfCRT,F145I mutation, associated with artemisinin and piperaquine resistance, respectively, a combination not found other isolates. All other isolates forming highly related clusters carried the K13,C580Y artemisinin resistance mutation. Core clusters were generally stable across IBD thresholds from 99% to 60%. Below 60%, clusters merged into larger components. Occupation, K13, and PfCRT mutations were consistently associated with cluster membership above 60% IBD (all p<0.05**; Supplementary Figure 2**). When isolates from Kratie were included, the collection province was strongly associated with cluster membership (p<0.007 all thresholds; **Supplementary Figure 3**).

**Figure 1.**
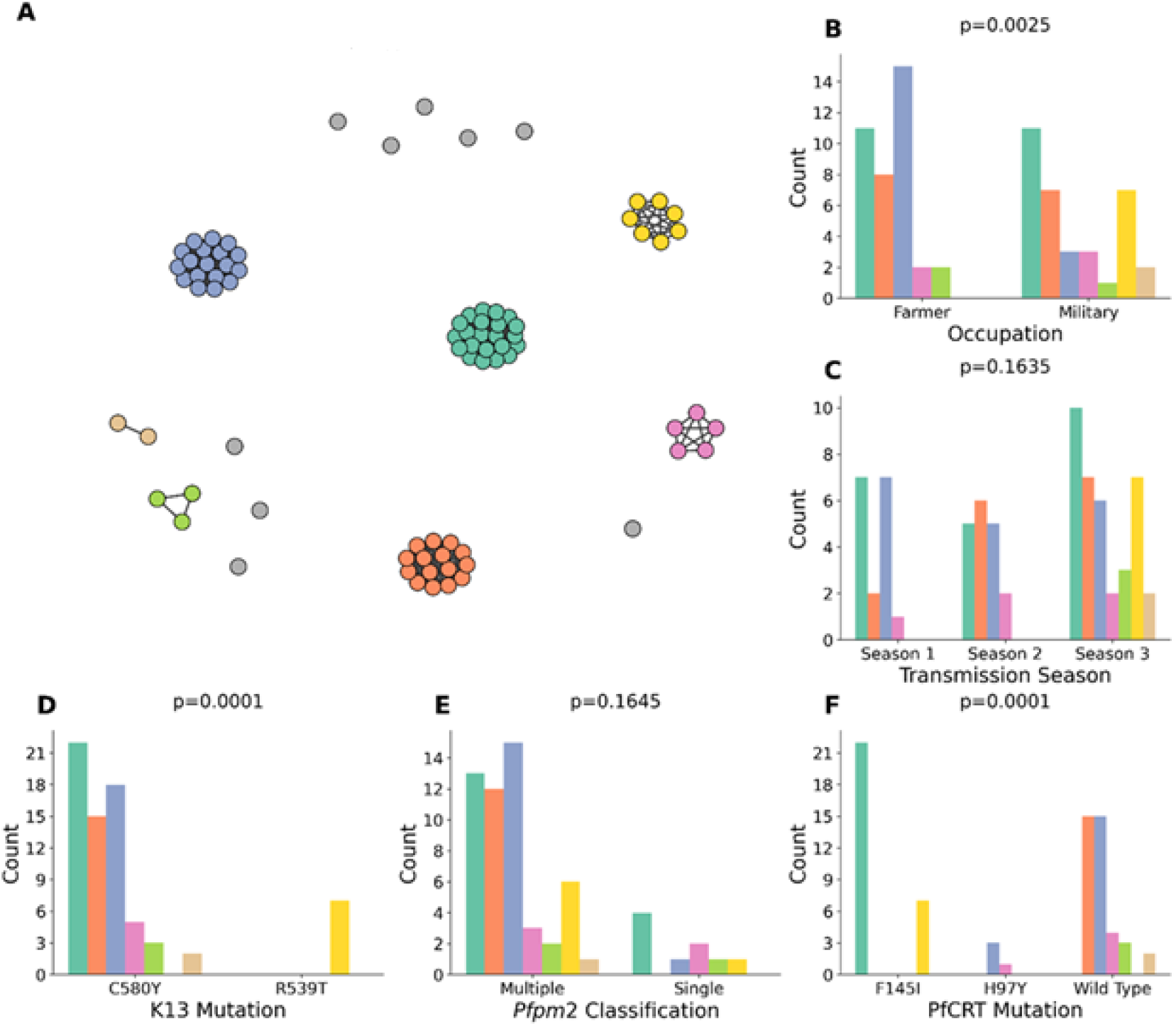
Highly Related Parasite Clusters and Associated Epidemiologic and Genetic Factors in Oddar Meanchey Province (0.90 IBD Threshold). Infomap network analysis identifies seven highly related parasite populations in Oddar Meanchey Province ranging from 2-22 members. **A**. The network graph shows parasite clusters, with colored nodes representing clonal populations and grey nodes representing unclustered isolates. **(B–F)** Bar plots show cluster membership among isolates in clusters with ≥2 isolates (panel A grey isolates excluded) by: **B**. Occupation, **C**. Transmission season, **D**. K13 resistance marker, **E**. *Pfpm2* copy number classification, and **F**. PfCRT mutation. Each bar color corresponds to a cluster. p-values were calculated using MCMC approximate Fisher’s exact tests.

### Association between predictors and highly related isolates

At the >90% threshold, isolate pairs from different occupations had 39% lower odds of being highly related than pairs from farmers, with strong posterior evidence (pd=0.958**; Table 3**). The wide CrI of the odds ratio (0.348–1.070), though predominantly below 1, likely reflects limited precision due to fewer pairs exceeding the >90% threshold. In contrast, isolates collected from military members had little posterior support for a difference in the odds of being highly related relative to farmers (pd=0.761). A one-year increase in age difference was associated with a small decrease in the odds of high relatedness (pd=0.966).

**Table 3.** Logistic regression compares the odds of high relatedness (>90% genome IBD) among different occupational groups controlling for difference in age.

|  | Odds Ratio | 95% CrI* | Pd** |
| --- | --- | --- | --- |
| **Occupation** |  |  |  |
| Farmer, Farmer | Ref (1.0) |  |  |
| Farmer, Military | 0.610 | (0.348, 1.070) | 0.958 |
| Military, Military | 0.682 | (0.236, 1.990) | 0.761 |
| **Age Difference (1 yr)** | 0.985 | (0.969, 1.001) | 0.966 |

\*Credible interval. \*\*Probability of direction.

Occupation remained a consistently associated across thresholds from 60–90% IBD. Across less stringent thresholds, CrIs were narrower and Pd values increased from the >90% IBD model, likely reflecting greater precision as more pairs met the high-relatedness definition (**Supplementary Tables 7D-G**). At >99% IBD, neither occupation contrast showed strong posterior support, likely due to fewer highly clonal pairs.

## Discussion

This study integrated genomic and epidemiologic data to assess how occupation relates to parasite genetic relatedness in Cambodian *P. falciparum* isolates. Isolate pairs from farmers shared more of their genome IBD than mixed-occupation pairs and had greater odds of being highly related, suggesting distinct sources of infection or exposure by occupation. Geographic location was the strongest predictor of relatedness, and the highest relatedness was observed between isolates in Oddar Meanchey. All parasites in highly related clusters harbored validated artemisinin-resistance markers, with R539T mutations present in a single military cluster, consistent with shared exposure within deployment or patrol groups. These findings underscore the importance of integrating genomic epidemiology into malaria surveillance, especially in low-transmission settings where factors like occupation may reflect both behavioral risk and unmeasured geographic heterogeneity in transmission. Leveraging IBD analysis alongside epidemiologic data can help refine control strategies, optimize targeted interventions, and improve understanding of transmission dynamics in pre-elimination contexts.

Circulating highly related *P. falciparum* lineages have been observed globally in low incidence settings.^15,40,41^ In this study, isolates from the same province were more closely related than to isolates from different provinces, in agreement with isolation-by-distance and previous reports of an inverse relationship between geographic distance and relatedness.^13,42^ Within-province relatedness was higher in Oddar Meanchey than Kratie. Isolates from Oddar Meanchey formed stable highly related clusters, consistent with prior reports of reduced genetic diversity following elimination efforts and the expansion of drug-resistant lineages. Previous temporal analyses of *P. falciparum* in Cambodia showed substantial increases in IBD from 2009 to 2014 and a lineage that rose to near fixation in Oddar Meanchey between 2014 and 2016.^14^

Within Oddar Meanchey, isolates from individuals with the same occupation were more closely related than those from different occupations, suggesting that farmers and military personnel are exposed to distinct transmission sources. These patterns indicate that geography is the primary driver of parasite relatedness, but occupation may further structure transmission patterns within geographic regions. Occupation may act as a proxy for differences in mobility patterns or time spent in forests, key malaria exposure determinants in this setting. In particular, military personnel are deployed to specific locations as part of their duties, while farmers’ exposure, tied to agricultural activities, may occur in different locations, leading to distinct patterns of forest contact between these groups. Occupation may also capture exposure-related behaviors like outdoor night work, net use, or housing type.^6^ Since fine-scale spatial data were unavailable, we could not fully disentangle geography from occupation, but the findings highlight how spatial and behavioral factors jointly shape transmission.

All parasites in highly related clusters carried validated markers of artemisinin resistance. Most had the K13,C580Y mutation, consistent with the expansion of highly related drug-resistant lineages in Cambodia during this period.^13,14,43^ PfCRT mutations, F145I and H97Y, were also associated with cluster membership. A single cluster of seven isolates all from military members carried K13 R539T and PfCRT F145I, a combination not observed in other parasites. While C580Y was the predominant artemisinin resistance mutation in Cambodia during this period, R539T was also present and observed along the Thai-Cambodian border, where military presence is high.^44–46^ The cluster of R539T infections among military personnel may reflect related infections acquired during deployment or patrol in forested areas, consistent with the fact these isolates were collected over a short time interval (January 26–February 10).

Participant age difference was associated with relatedness, with more similar-aged individuals harboring more closely related parasites. The effect size was small, but this association could reflect shared exposures or behaviors among similarly aged individuals, such as co-travel, co-residence, or occupation-linked activities not captured in the metadata. Farmers were on average younger, which may contribute to age-related structure in parasite populations if exposure differs by occupation. In contrast, the time between sample collection dates was not associated with genetic relatedness, despite expectation that samples collected closer together in time may be more related. This lack of association likely reflects the short sampling window (<2 years) and persistence of resistant lineages in a low-transmission setting with limited recombination.^14,47^

Although this study demonstrates the value of integrating genomic and epidemiologic data, its interpretation was limited by the scope and granularity of available covariates. The baseline beta regression model with a multiple membership random effects already explained ∼16.7% of the variation in parasite relatedness, reflecting the strong influence of repeated appearance of isolates in pairs. Adding occupation and age improved the model fit modestly (R^2^=0.172), indicating that the measured predictors explained a relatively small additional fraction of the variation. Most variation likely reflects unmeasured geographic, environmental, or host factors. Our dataset was drawn from two separate clinical studies, and although occupation’s effect persisted after adjusting for study source, residual differences in study design may have influenced results. Additionally, one study used a cluster-randomized design, but cluster locations were unavailable for security reasons due to military participation, preventing adjustment for within-cluster correlation or testing whether occupation partly reflects geographic clustering. Occupation may therefore serve as a proxy for unmeasured spatial factors. Finer-grained location data could have improved adjustment for spatial confounding. Additional metadata on household relationships or travel histories would further enhance inference. Recent work in Cambodia has shown that forest-going activities are frequently misclassified as farming in epidemiologic studies, potentially introducing misclassification bias.^48^ The result would likely be a more heterogeneous farmer group and less related parasites on average, biasing results toward the null. Because detailed occupational sub-categories were not collected, the farmer category in our dataset may reflect a mix of agricultural and forest-based activities, limiting our ability to distinguish exposure differences within this group.

These results suggest that farmers and military personnel may acquire infections from different exposure settings and benefit from tailored interventions. Intermittent preventive treatment, involving periodic administration of antimalarials regardless of symptoms, has been effective in hard-to-reach populations and could be adapted for mobile farmers during harvest seasons when care access is limited.^48^ In contrast, monthly malaria prophylaxis offers continuous protection and is effective in military settings, where centralized health infrastructure supports regular dosing and follow-up.^19^ Occupation-specific strategies may be necessary because behavioral and structural factors shape infection risk and exposure context. For example, farmers may sleep in open or partially enclosed field shelters during harvest season, facing greater risk of night-time mosquito bites.^49^ Farmers and other civilian forest-goers may also miss intervention campaigns if they are away in forests or fields during distribution, or delay seeking treatment due to economic or geographic barriers.^50^ These barriers are potentially less relevant for military personnel with centralized healthcare access. Our results demonstrate the value of combining genomic and epidemiologic data to improve transmission inference and guide targeted interventions in malaria elimination settings.

## Data Availability

All data produced in the present study are available upon reasonable request to the authors. All sequence data are publicly available (NCBI SRA access number PRJNA1004408).

## Disclaimer

Material has been reviewed by the Walter Reed Army Institute of Research. There is no objection to its presentation and/or publication. The opinions or assertions contained herein are the private views of the author, and are not to be construed as official, or as reflecting true views of the Department of the Army or the Department of Defense. The investigators have adhered to the policies for protection of human subjects as prescribed in AR 70–25.

## Acknowledgements

We thank all participants in the original studies. We thank Melissa Meyers for program support. We thank Jean Paul Courneya for data management assistance and the Malaria Research Program Laboratory Staff for sample processing and management.

## Contributions

Conceptualization: E.R., M.W., J.S., T.O., S.T.H; Formal Analysis: E.R.; Data Curation: B.G., M.W; Investigation: M.S., B.V., S.Sri., S.P., S.C., S.Som., S.T., P.S., C.C.; Project Administration: M.S., S.Sri., D.S., D.L., C.L., P.L.; Software: B.G.; Funding Acquisition: T.O., S.T.H.; Supervision: N.W., D.S., S.T.H Writing – original draft: E.R.; Writing – review & editing: B.V, M.S, D.S., S.T.H

