## Supplementary Figure 1 for "Genetic relatedness of Cambodian *Plasmodium falciparum* isolates is driven by geography and occupation"

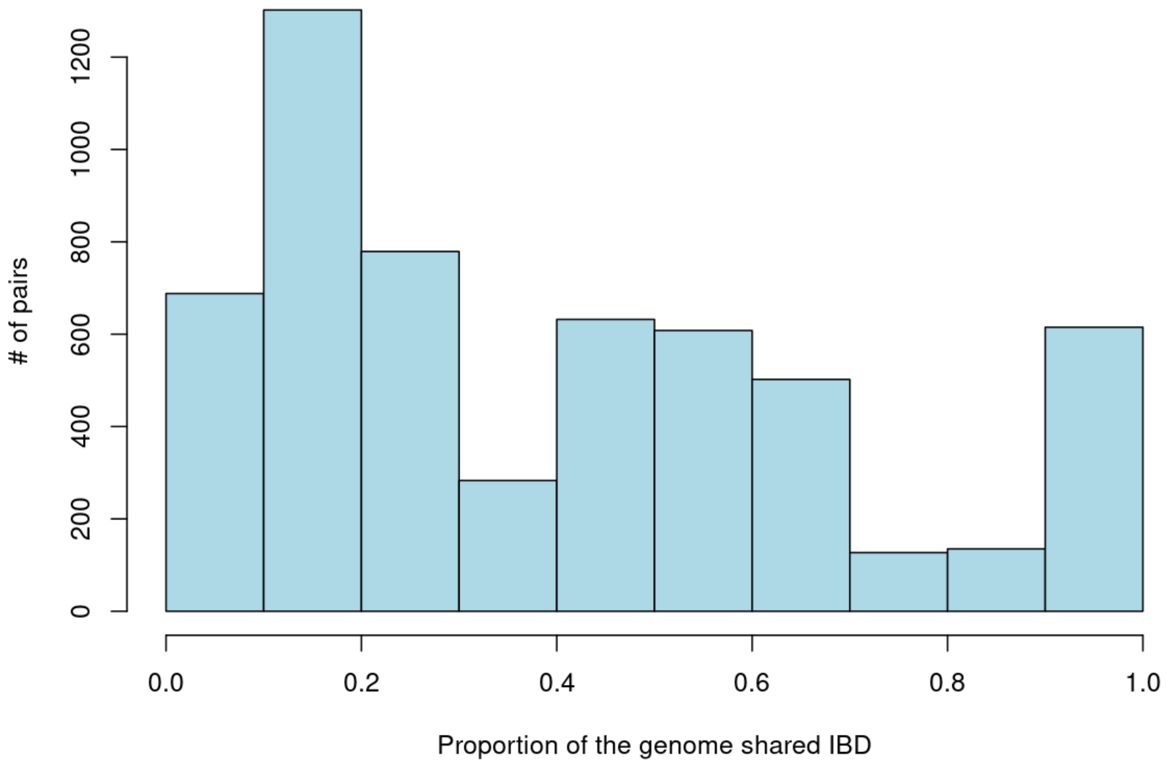


**Supplementary Figure 1.** Distribution of pairwise identity-by-descent (IBD) across all isolate pairs. The histogram shows a multimodal distribution with peaks at low (<0.2), moderate (0.4–0.7), and clonal (>0.9) relatedness levels.
