## Supplementary Figure 2 for "Genetic relatedness of Cambodian *Plasmodium falciparum* isolates is driven by geography and occupation"

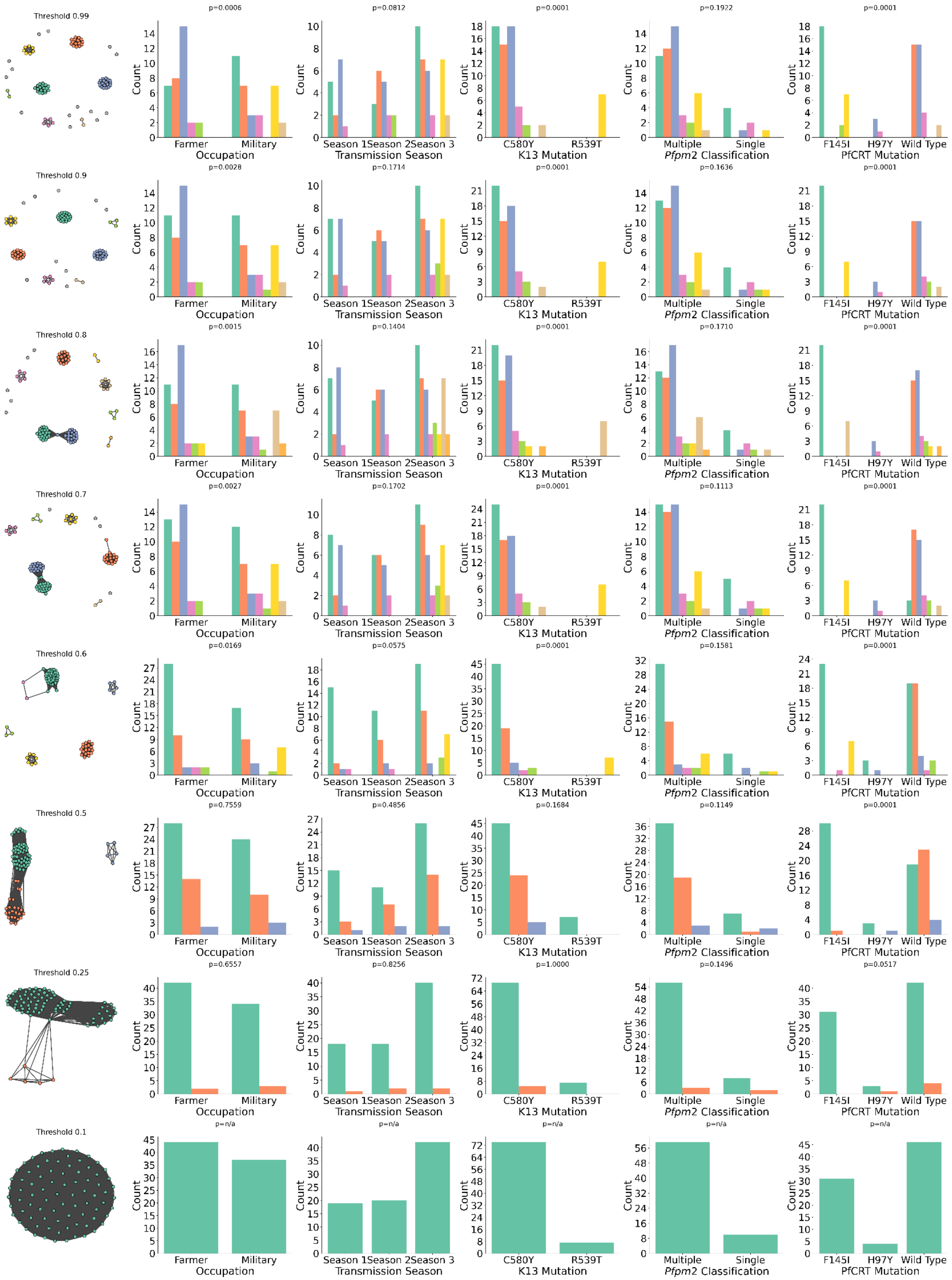
Threshold 0.99
p=0.0006
p=0.0812
p=0.0001
p=0.1922
p=0.0001
Threshold 0.9
p=0.0028
p=0.1714
p=0.0001
p=0.1636
p=0.0001
Threshold 0.8
p=0.0015
p=0.1404
p=0.0001
p=0.1710
p=0.0001
Threshold 0.7
p=0.0027
p=0.1702
p=0.0001
p=0.1113
p=0.0001
Threshold 0.6
p=0.0169
p=0.0575
p=0.0001
p=0.1581
p=0.0001
Threshold 0.5
p=0.7559
p=0.4856
p=0.1684
p=0.1149
p=0.0001
Threshold 0.25
p=0.6557
p=0.8256
p=1.0000
p=0.1496
p=0.0517
Threshold 0.1
p=n/a
p=n/a
p=n/a
p=n/a
p=n/a
Count
Farmer
Military
Occupation
Season 1
Season 2
Season 3
Transmission Season
C580Y
R539T
K13 Mutation
Multiple
Single
*Pfpm2* Classification
F145I
H97Y
Wild Type
PfCRT Mutation

**Supplementary Figure 2.** Network-Based Analysis of Parasite Clustering Across Relatedness Thresholds and Associated Epidemiologic and Genetic Factors in Oddar Meanchey Province. This figure illustrates parasite clustering and the association of cluster composition with epidemiologic and genetic variables across varying pairwise relatedness thresholds (0.99 to 0.1). Network graphs depict parasite relatedness networks at each threshold, where nodes represent individual parasite isolates and edges indicate pairwise genetic relatedness above the specified threshold. Distinct colors indicate clusters identified using the Infomap algorithm. As thresholds decrease, networks become more interconnected, leading to the formation of larger clusters. Bar plots show the distribution of epidemiologic factors (Occupation and Transmission Season) and genetic factors (K13 Mutation, *Pfpm2* Copy Number Classification, PfCRT Mutation) within identified clusters at each threshold. Each bar color corresponds to a cluster from the network graphs. p-values were calculated using MCMC approximate Fisher's exact tests to assess the strength of association between cluster membership and each variable.
