## Supplementary Table 1 for "Genetic relatedness of Cambodian *Plasmodium falciparum* isolates is driven by geography and occupation"

**Supplementary Table 1.** Metadata of the population included in IBD analyses (N=107). Submission IDs are anonymized IDs assigned to the parasite sequences and are not patient study IDs (NCBI SRA access number PRJNA1004408).

| Submission ID | Occupation | Year | Province* | Collection Month** | Age Range** | Sex | Polyclonal By Fws | Symptom Status |
|---|---|---|---|---|---|---|---|---|
| CVD_KHM_227 | Farmer | 2014 | OM | December | 18-30 | Male | Yes | Yes |
| CVD_KHM_228 | Farmer | 2014 | OM | December | 18-30 | Male | Yes | Yes |
| CVD_KHM_229 | Farmer | 2014 | OM | December | 18-30 | Male | No | Yes |
| CVD_KHM_230 | Farmer | 2014 | OM | December | 31-40 | Male | Yes | Yes |
| CVD_KHM_231 | Farmer | 2014 | OM | December | 18-30 | Male | No | Yes |
| CVD_KHM_232 | Farmer | 2014 | OM | December | 31-40 | Female | No | Yes |
| CVD_KHM_233 | Farmer | 2014 | OM | December | 41+ | Male | No | Yes |
| CVD_KHM_273 | Farmer | 2015 | OM | January | 18-30 | Male | No | Yes |
| CVD_KHM_274 | Farmer | 2015 | OM | January | 31-40 | Male | No | Yes |
| CVD_KHM_275 | Farmer | 2015 | OM | January | 31-40 | Male | No | Yes |
| CVD_KHM_276 | Farmer | 2015 | OM | January | 18-30 | Male | No | Yes |
| CVD_KHM_279 | Farmer | 2015 | OM | January | 18-30 | Male | No | Yes |
| CVD_KHM_280 | Farmer | 2015 | OM | January | 18-30 | Male | No | Yes |
| CVD_KHM_281 | Farmer | 2015 | OM | January | 18-30 | Male | No | Yes |
| CVD_KHM_282 | Military | 2015 | OM | January | 31-40 | Male | No | Yes |
| CVD_KHM_283 | Farmer | 2015 | OM | February | 18-30 | Male | No | Yes |
| CVD_KHM_284 | Farmer | 2015 | OM | February | 31-40 | Male | No | Yes |
| CVD_KHM_285 | Farmer | 2015 | OM | February | 41+ | Male | No | Yes |
| CVD_KHM_286 | Farmer | 2015 | OM | February | 41+ | Male | No | Yes |
| CVD_KHM_288 | Farmer | 2015 | OM | April | 18-30 | Male | No | Yes |
| CVD_KHM_289 | Farmer | 2015 | OM | May | 31-40 | Male | No | Yes |
| CVD_KHM_290 | Farmer | 2015 | OM | June | 18-30 | Male | No | Yes |
| CVD_KHM_291 | Farmer | 2015 | OM | June | 18-30 | Male | No | Yes |
| CVD_KHM_293 | Farmer | 2015 | OM | June | 18-30 | Male | No | Yes |
| CVD_KHM_294 | Farmer | 2015 | OM | June | 41+ | Male | No | Yes |
| CVD_KHM_296 | Military | 2015 | OM | July | 41+ | Male | No | Yes |
| CVD_KHM_297 | Farmer | 2015 | OM | July | 18-30 | Male | No | Yes |
| CVD_KHM_299 | Farmer | 2015 | OM | July | 31-40 | Male | No | Yes |
| CVD_KHM_300 | Farmer | 2015 | OM | July | 31-40 | Female | No | Yes |
| CVD_KHM_301 | Farmer | 2015 | OM | July | 18-30 | Male | No | Yes |
| CVD_KHM_302 | Farmer | 2015 | OM | July | 31-40 | Male | No | Yes |
| CVD_KHM_303 | Farmer | 2015 | OM | July | 18-30 | Male | No | Yes |
| CVD_KHM_304 | Farmer | 2015 | OM | July | 18-30 | Male | No | Yes |
| CVD_KHM_305 | Farmer | 2015 | OM | July | 18-30 | Male | No | Yes |
| CVD_KHM_306 | Farmer | 2015 | OM | July | 18-30 | Male | No | Yes |
| CVD_KHM_307 | Farmer | 2015 | OM | July | 18-30 | Male | No | Yes |
| CVD_KHM_308 | Farmer | 2015 | OM | August | 31-40 | Male | Yes | Yes |
| CVD_KHM_311 | Farmer | 2015 | OM | September | 31-40 | Male | No | Yes |

| | | | | | | | | |
|---|---|---|---|---|---|---|---|---|
| CVD_KHM_312 | Farmer | 2015 | OM | September | 18-30 | Male | No | Yes |
| CVD_KHM_313 | Farmer | 2015 | KT | May | 41+ | Male | No | Yes |
| CVD_KHM_315 | Farmer | 2015 | KT | May | 41+ | Male | No | Yes |
| CVD_KHM_316 | Farmer | 2015 | KT | May | 18-30 | Male | Yes | Yes |
| CVD_KHM_317 | Farmer | 2015 | KT | May | 18-30 | Male | No | Yes |
| CVD_KHM_318 | Farmer | 2015 | KT | June | 18-30 | Male | No | Yes |
| CVD_KHM_319 | Farmer | 2015 | KT | June | 41+ | Male | No | Yes |
| CVD_KHM_320 | Farmer | 2015 | KT | June | 18-30 | Male | No | Yes |
| CVD_KHM_321 | Farmer | 2015 | KT | June | 41+ | Male | No | Yes |
| CVD_KHM_322 | Farmer | 2015 | KT | July | 18-30 | Male | No | Yes |
| CVD_KHM_323 | Farmer | 2015 | KT | July | 31-40 | Male | No | Yes |
| CVD_KHM_324 | Farmer | 2015 | KT | July | 31-40 | Male | No | Yes |
| CVD_KHM_326 | Farmer | 2015 | KT | August | 18-30 | Male | No | Yes |
| CVD_KHM_327 | Farmer | 2015 | KT | August | 18-30 | Male | No | Yes |
| CVD_KHM_328 | Farmer | 2015 | KT | August | 18-30 | Male | No | Yes |
| CVD_KHM_329 | Farmer | 2015 | KT | August | 18-30 | Male | No | Yes |
| CVD_KHM_330 | Farmer | 2015 | KT | August | 41+ | Male | Yes | Yes |
| CVD_KHM_331 | Farmer | 2015 | KT | August | 31-40 | Male | No | Yes |
| CVD_KHM_332 | Farmer | 2015 | KT | August | 41+ | Male | No | Yes |
| CVD_KHM_333 | Farmer | 2015 | KT | September | 31-40 | Male | No | Yes |
| CVD_KHM_334 | Farmer | 2015 | KT | September | 18-30 | Male | Yes | Yes |
| CVD_KHM_335 | Farmer | 2015 | KT | September | 31-40 | Male | No | Yes |
| CVD_KHM_336 | Farmer | 2015 | KT | September | 18-30 | Male | No | Yes |
| CVD_KHM_337 | Farmer | 2015 | KT | September | 18-30 | Male | No | Yes |
| CVD_KHM_339 | Farmer | 2015 | KT | September | 18-30 | Male | No | Yes |
| CVD_KHM_340 | Farmer | 2015 | KT | September | 18-30 | Male | No | Yes |
| CVD_KHM_341 | Farmer | 2015 | KT | September | 31-40 | Male | No | Yes |
| CVD_KHM_342 | Military | 2016 | OM | January | 31-40 | Male | No | Yes |
| CVD_KHM_343 | Military | 2016 | OM | January | 41+ | Male | No | Yes |
| CVD_KHM_344 | Military | 2016 | OM | January | 18-30 | Male | No | No |
| CVD_KHM_345 | Military | 2016 | OM | January | 41+ | Male | No | Yes |
| CVD_KHM_346 | Military | 2016 | OM | January | 41+ | Male | No | No |
| CVD_KHM_347 | Military | 2016 | OM | January | 41+ | Male | No | Yes |
| CVD_KHM_349 | Military | 2016 | OM | March | 31-40 | Male | No | Yes |
| CVD_KHM_351 | Military | 2016 | OM | February | 31-40 | Male | No | No |
| CVD_KHM_352 | Farmer | 2016 | OM | January | 20-30 | Male | No | Yes |
| CVD_KHM_353 | Farmer | 2016 | OM | January | 18-30 | Male | No | No |
| CVD_KHM_354 | Farmer | 2016 | OM | January | 41+ | Female | No | No |
| CVD_KHM_355 | Farmer | 2016 | OM | February | 31-40 | Male | No | No |
| CVD_KHM_357 | Farmer | 2016 | OM | January | <18 | Female | Yes | No |
| CVD_KHM_358 | Farmer | 2016 | OM | June | 41+ | Female | No | Yes |
| CVD_KHM_359 | Farmer | 2016 | OM | January | 41+ | Male | No | Yes |
| CVD_KHM_360 | Military | 2016 | OM | January | 41+ | Male | No | Yes |

| | | | | | | | | |
|---|---|---|---|---|---|---|---|---|
| CVD_KHM_361 | Military | 2016 | OM | January | 31-40 | Male | No | Yes |
| CVD_KHM_362 | Military | 2016 | OM | February | 18-30 | Male | Yes | No |
| CVD_KHM_366 | Military | 2016 | OM | January | 41+ | Male | No | Yes |
| CVD_KHM_367 | Military | 2016 | OM | January | 31-40 | Male | No | Yes |
| CVD_KHM_368 | Military | 2016 | OM | January | 31-40 | Male | Yes | No |
| CVD_KHM_369 | Military | 2016 | OM | January | 31-40 | Male | No | Yes |
| CVD_KHM_370 | Military | 2016 | OM | January | 31-40 | Male | No | Yes |
| CVD_KHM_371 | Military | 2016 | OM | January | 31-40 | Male | No | Yes |
| CVD_KHM_372 | Military | 2016 | OM | January | 18-30 | Male | Yes | No |
| CVD_KHM_373 | Military | 2016 | OM | January | 31-40 | Male | No | Yes |
| CVD_KHM_375 | Military | 2016 | OM | January | 18-30 | Male | No | No |
| CVD_KHM_376 | Military | 2016 | OM | January | 41+ | Male | No | Yes |
| CVD_KHM_377 | Military | 2016 | OM | January | 18-30 | Male | No | Yes |
| CVD_KHM_378 | Military | 2016 | OM | February | 18-30 | Male | No | Yes |
| CVD_KHM_379 | Military | 2016 | OM | February | 18-30 | Male | No | Yes |
| CVD_KHM_380 | Military | 2016 | OM | February | 18-30 | Male | No | No |
| CVD_KHM_381 | Military | 2016 | OM | February | 31-40 | Male | No | No |
| CVD_KHM_382 | Military | 2016 | OM | February | 31-40 | Male | No | Yes |
| CVD_KHM_383 | Military | 2016 | OM | February | 41+ | Male | No | No |
| CVD_KHM_384 | Military | 2016 | OM | February | 31-40 | Male | No | No |
| CVD_KHM_385 | Military | 2016 | OM | February | 41+ | Male | No | Yes |
| CVD_KHM_387 | Military | 2016 | OM | February | 18-30 | Male | No | Yes |
| CVD_KHM_388 | Military | 2016 | OM | February | 18-30 | Male | No | Yes |
| CVD_KHM_389 | Military | 2016 | OM | February | 31-40 | Male | No | Yes |
| CVD_KHM_390 | Military | 2016 | OM | February | 41+ | Male | No | Yes |
| CVD_KHM_391 | Military | 2016 | OM | February | 31-40 | Male | No | No |

* OM is Oddar Meanchey province and KT is Kratie province. ** Collection date and age are provided in ranges to preserve participant anonymity.
