## Supplementary Table 2 for "Genetic relatedness of Cambodian *Plasmodium falciparum* isolates is driven by geography and occupation"

**Supplementary Table 2.** Descriptive categorical characteristics of the population included in IBD analyses (N=107).

| Covariate | n (%) |
|---|---|
| **Occupation** | |
| Farmer | 37 (35.0) |
| Military | 70 (65.0) |
| **Study** | |
| Study 1 | 65 (60.7) |
| Study 2 | 42 (39.3) |
| **Symptom Status** | |
| Symptomatic | 93 (86.9) |
| Asymptomatic | 14 (13.1) |
| **Province** | |
| Kratie | 26 (24.0) |
| Oddar Meanchey | 81 (76.0) |
| **Sex** | |
| Male | 102 (95.0) |
| Female | 5 (5.0) |
| **Season*** | |
| Season 1 | 19 (18.0) |
| Season 2 | 46 (43.0) |
| Season 3 | 42 (39.0) |
| **Age** | 33.6 (10.0)** |

* Season 1 was from December 2014 to April 15th, 2015. Season 2 was April 16th, 2015, to September 2015 and Season 3 (January 2016 – June 2016). ** Age variable is reported as mean (standard deviation).
