## Supplementary Table 3 for "Genetic relatedness of Cambodian *Plasmodium falciparum* isolates is driven by geography and occupation"

**Supplementary Table 3.** Descriptive categorical characteristics of the study population (N=313).

| Covariate | n (%) |
|---|---|
| **Occupation** | |
| Farmer | 205 (65.5) |
| Military | 99 (31.6) |
| Other | 9 (2.9) |
| **Study** | |
| Study 1 | 205 (65.5) |
| Study 2 | 108 (34.5) |
| **Province** | |
| Kratie | 48 (15.3) |
| Oddar Meanchey | 265 (84.7) |
| **Sex** | |
| Male | 301 (96.2) |
| Female | 12 (3.8) |
| **Season*** | |
| Season 1 | 41 (13.1) |
| Season 2 | 164 (52.4) |
| Season 3 | 109 (34.8) |
| **Age** | 30.3 (16.4)** |

* Season 1 was from December 2014 to April 15th, 2015. Season 2 was April 16th, 2015, to September 2015 and Season 3 (January 2016 – June 2016). ** Age variable is reported at mean (standard deviation).
