## Supplementary Table 4 for "Genetic relatedness of Cambodian *Plasmodium falciparum* isolates is driven by geography and occupation"

**Supplementary Table 4.** Descriptive characteristics of study population by occupational pair grouping.

| **Covariate** | **Occupation n (%)** | | | Total | p-value |
|---|---|---|---|---|---|
| | Different Occupation 2590 (46) | Both Farmers 2415 (42) | Both Military 666 (12) | 5671(100) | |
| **Collection Province** | | | | | <2.2e-16** |
| KT, KT | 0 (0) | 325 (100) | 0 (0) | 325 (100) | |
| OM, KT | 962 (46) | 1144 (54) | 0 (0) | 2106 (100) | |
| OM, OM | 1628 (50) | 946 (29) | 666 (21) | 3240 (100) | |
| **Sex** | | | | | < 2.2e-16ᵀ |
| Female, Female | 0 (0) | 10 (100) | 0 (0) | 10 (100) | |
| Female, Male | 185 (36) | 325 (64) | 0 (0) | 510 (100) | |
| Male, Male | 2405 (47) | 2080 (40) | 666 (13) | 5151 (100) | |
| **Days between collection** (mean, 95% CI) | 224.2 (220, 229) | 131.9 (128, 136) | 50.2 (43, 57) | 164.4 (161, 167) | < 2.2e-16^ |
| **Age difference** (mean, 95% CI) | 11.7 (11.3, 12.0) | 11.3 (11.0, 11.7) | 10.4 (9.9, 11.0) | 11.4 (11.2, 11.6) | 0.013^ |

* Percentages displayed indicate percentage across rows. ** p-value calculated using chi-square test. ᵀ p-value calculated using Fisher's exact test. ^ p-values calculated using Kruskal-Wallis test.
