## Supplementary Table 5 for "Genetic relatedness of Cambodian *Plasmodium falciparum* isolates is driven by geography and occupation"

**Supplementary Table 5.** Descriptive characteristics of study population in Oddar Meanchey province (N = 3,240)

| | n (%) | Mean IBD | 95% CI | p-value |
|---|---|---|---|---|
| **Occupation** | | | | 5.74e-12* |
| Farmer, Farmer | 946 (29.2) | 0.538 | (0.519, 0.558) | |
| Farmer, Military | 1628 (50.2) | 0.472 | (0.458, 0.486) | |
| Military, Military | 666 (20.6) | 0.451 | (0.428, 0.473) | |
| **Sex** | | | | |
| Male, Male | 2850 (88.0) | 0.481 | (0.470, 0.491) | 1.68e-04* |
| Female, Male | 380 (11.7) | 0.533 | (0.506, 0.559) | |
| Female, Female | 10 (0.3) | 0.544 | (0.432, 0.658) | |
| **Days between collection** | 3240 (100) | 180.4 | (175.4, 185.4) | 0.485^ |
| **Age difference (years)** | 3240 (100) | 11.3 | (11.1, 11.6) | 0.008^ |

* p-values calculated based on Kruskal-Wallis test ^ p-values calculated based on Spearman's correlation coefficient
