## Supplementary Table 6 for "Genetic relatedness of Cambodian *Plasmodium falciparum* isolates is driven by geography and occupation"

**Supplementary Table 6A.** Baseline beta regression model with multiple membership random effects only (no fixed predictors). $R^2 = 0.167$.

| | Parameter estimates | Marginal change in proportion IBD | 95% CI | Probability of direction (pd) |
|---|---|---|---|---|
| **Random effects** | 0.51 | 0.624 | (0.580, 0.666) | |

* phi = 1.06 (1.01, 1.10)

**Supplementary Table 6B.** Beta regression comparing the proportion of the genome shared IBD between occupation groups. $R^2 = 0.171$.

| | Parameter estimates | Marginal change in proportion IBD | 95% CI | Probability of direction (pd) |
|---|---|---|---|---|
| **Occupation** | | | | |
| Famer, Farmer | 0.70 | 0.668 | (0.610, 0.722) | |
| Military, Famer | -0.31 | -0.071 | (-0.116, -0.025) | 99.9 |
| Military, Military | -0.22 | -0.050 | (-0.135, 0.038) | 87.2 |

* phi = 1.06 (1.02, 1.11)

**Supplementary Table 6C.** Beta regression comparing the proportion of the genome shared IBD between occupation groups adjusted for difference in collection days. $R^2 = 0.172$.

| | Parameter estimates | Marginal change in proportion IBD | 95% CI | Probability of direction (pd) |
|---|---|---|---|---|
| **Occupation** | | | | |
| Famer, Farmer | 0.68 | 0.670 | (0.611, 0.723) | |
| Military, Famer | -0.32 | -0.074 | (-0.121, -0.026) | 99.9 |
| Military, Military | -0.20 | -0.047 | (-0.135, 0.040) | 85.0 |
| **One day difference in collection** | 0.00 | 0.000 | (0.000, 0.000) | 77.4 |

*phi = 1.06 (1.02, 1.11)

**Supplementary Table 6D.** Beta regression comparing the proportion of the genome shared IBD between pairs from the same or different collection studies. $R^2 = 0.168$

| | Parameter estimates | Marginal change in proportion IBD | 95% CI | Probability of direction (pd) |
|---|---|---|---|---|
| **Study** | | | | |
| Same Study | 0.55 | 0.633 | (0.589, 0.677) | |
| Different Study | -0.08 | -0.018 | (-0.038, 0.002) | 96.2 |

* phi = 1.06 (1.01, 1.10)

**Supplementary Table 6E.** Beta regression comparing the proportion of the genome shared IBD between occupation groups controlling for difference in collection study. $R^2 = 0.172$

| | Parameter estimates | Marginal change in proportion IBD | 95% CI | Probability of direction (pd) |
|---|---|---|---|---|
| **Occupation** | | | | |
| Famer, Farmer | 0.68 | 0.664 | (0.605, 0.720) | |

| | | | | |
|---|---|---|---|---|
| Military, Famer | -0.35 | -0.079 | (-0.126, -0.033) | 99.9 |
| Military, Military | -0.21 | -0.047 | (-0.131, 0.036) | 86.2 |
| **Study** | | | | |
| Different Study | 0.07 | 0.0160 | (-0.009 , 0.041) | 89.4 |

* phi = 1.06 (1.02, 1.11)

**Supplementary Table 6F.** Beta regression comparing the proportion of the genome shared IBD between symptomatic and asymptomatic cases. $R^2 = 0.168$

| | Parameter estimates | Marginal change in proportion IBD | 95% CI | Probability of direction (pd) |
|---|---|---|---|---|
| **Symptom status** | | | | |
| Both asymptomatic | 0.33 | 0.581 | (0.468, 0.688) | |
| One symptomatic | 0.15 | 0.036 | (-0.040, 0.118) | 81.2 |
| Both symptomatic | 0.20 | 0.047 | (-0.068 , 0.171) | 78.0 |

* phi = 1.06 (1.02, 1.11)

**Supplementary Table 6G.** Beta regression comparing the proportion of the genome shared IBD between occupation groups controlling for case symptom status. $R^2 = 0.172$

| | Parameter estimates | Marginal change in proportion IBD | 95% CI | Probability of direction (pd) |
|---|---|---|---|---|
| **Occupation** | | | | |
| Famer, Farmer | 0.57 | 0.637 | (0.503, 0.755) | |
| Military, Famer | -0.31 | -0.071 | (-0.244, 0.098) | 99.9 |
| Military, Military | -0.21 | -0.048 | (-0.219, 0.120) | 86.2 |
| **Symptom status** | | | | |
| Both asymptomatic | 0.57 | 0.637 | (0.503 , 0.755) | |
| One symptomatic | 0.14 | 0.035 | (-0.042, 0.118) | 79.7 |
| Both symptomatic | 0.14 | 0.033 | (-0.087, 0.160) | 69.8 |

* phi = 1.06 (1.02, 1.11)
