## Supplementary Table 7 for "Genetic relatedness of Cambodian *Plasmodium falciparum* isolates is driven by geography and occupation"

**Supplementary Table 7A.** Logistic regression compares the odds of high relatedness >10% genome IBD among different occupational groups controlling for collection province and age difference**.**

| | Odds Ratio | 95% CI | Probability of Direction |
|---|---|---|---|
| **Occupation** | | | |
| Famer, Farmer | Ref (1.0) | | 1 |
| Farmer, Military | Inf | (0.000, Inf) | 0.596 |
| Military, Military | 6.411E+29 | (0.000, Inf) | 0.570 |
| **Age Difference (1 yr)** | 3.506E+127 | (0.000, Inf) | 0.711 |

**Supplementary Table 7B.** Logistic regression compares the odds of high relatedness >25% genome IBD among different occupational groups controlling for collection province and age difference**.**

| | Odds Ratio | 95% CI | Probability of Direction |
|---|---|---|---|
| **Occupation** | | | |
| Famer, Farmer | Ref (1.0) | | |
| Farmer, Military | 0.797 | (0.322, 1.940) | 0.687 |
| Military, Military | 0.523 | (0.089, 2.977) | 0.762 |
| **Age Difference (1 yr)** | 0.996 | (0.979, 1.013) | 0.678 |

**Supplementary Table 7C.** Logistic regression compares the odds of high relatedness >50% genome IBD among different occupational groups controlling for collection province and age difference**.**

| | Odds Ratio | 95% CI | Probability of Direction |
|---|---|---|---|
| **Occupation** | | | |
| Famer, Farmer | Ref (1.0) | | |
| Farmer, Military | 0.702 | (0.426, 1.143) | 0.921 |
| Military, Military | 0.623 | (0.242, 1.573) | 0.840 |
| **Age Difference (1 yr)** | 0.997 | (0.984, 1.010) | 0.685 |

**Supplementary Table 7D.** Logistic regression compares the odds of high relatedness >60% genome IBD among different occupational groups controlling for collection province and age difference**.**

| | Odds Ratio | 95% CI | Probability of Direction |
|---|---|---|---|
| **Occupation** | | | |
| Famer, Farmer | Ref (1.0) | | |
| Farmer, Military | 0.468 | (0.245, 0.912) | 0.986 |
| Military, Military | 0.273 | (0.078, 0.997) | 0.975 |
| **Age Difference (1 yr)** | 0.984 | (0.9702, 0.999) | 0.983 |

**Supplementary Table 7E.** Logistic regression compares the odds of high relatedness >70% genome IBD among different occupational groups controlling for collection province and age difference**.**

| | Odds Ratio | 95% CI | Probability of Direction |
|---|---|---|---|
| **Occupation** | | | |
| Famer, Farmer | Ref (1.0) | | |
| Farmer, Military | 0.435 | (0.260, -0.729) | 0.999 |
| Military, Military | 0.419 | (0.160, 1.13) | 0.957 |
| **Age Difference (1 yr)** | 0.992 | (0.977, 1.007) | 0.861 |

**Supplementary Table 7F.** Logistic regression compares the odds of high relatedness >80% genome IBD among different occupational groups controlling for collection province and age difference**.**

| | Odds Ratio | 95% CI | Probability of Direction |
|---|---|---|---|
| **Occupation** | | | |
| Famer, Farmer | Ref (1.0) | | |
| Farmer, Military | 0.461 | (0.271, 0.779) | 0.998 |
| Military, Military | 0.447 | (0.1635, 1.200) | 0.946 |
| **Age Difference (1 yr)** | 0.989 | (0.974, 1.004) | 0.915 |

**Supplementary Table 7G.** Logistic regression compares the odds of high relatedness >90% genome IBD among different occupational groups controlling for collection province and age difference**.**

| | Odds Ratio | 95% CI | Probability of Direction |
|---|---|---|---|
| **Occupation** | | | |
| Famer, Farmer | Ref (1.0) | | |
| Farmer, Military | 0.6104 | (0.348, 1.070) | 0.958 |
| Military, Military | 0.682 | (0.235, 1.990) | 0.761 |
| **Age Difference (1 yr)** | 0.985 | (0.969, -1.001) | 0.966 |

**Supplementary Table 7H.** Logistic regression compares the odds of high relatedness >99% genome IBD among different occupational groups controlling for collection province and age difference**.**

| | Odds Ratio | 95% CI | Probability of Direction |
|---|---|---|---|
| **Occupation** | | | |
| Famer, Farmer | Ref (1.0) | | |
| Farmer, Military | 0.692 | (0.361, 1.358) | 0.864 |
| Military, Military | 1.235 | (0.357, 4.521) | 0.630 |
| **Age Difference (1 yr)** | 0.992 | (0.975, 1.010) | 0.806 |
